# Forecasting the COVID-19 Pandemic with Climate Variables for Top Five Burdening and Three South Asian Countries

**DOI:** 10.1101/2020.05.12.20099044

**Authors:** Md. Karimuzzaman, Sabrina Afroz, Md. Moyazzem Hossain, Azizur Rahman

**Affiliations:** Department of Statistics, Jahangirnagar University, Savar, Dhaka, Bangladesh; School of Mathematics, Statistics and Physics, Newcastle University, Newcastle, UK; School of Computing and Mathematics, Charles Sturt University, Wagga Wagga, Australia

**Keywords:** COVID-19, Climate Variables, Count Time Series, likelihood based GLM, Machine Learning

## Abstract

**Background:** The novel coronavirus (COVID-19) is now in a horrific situation around the world. Prediction about the number of infected and death cases may help to take immediate action to prevent the epidemic as well as control the situation of a country. The ongoing debate about the climate factors may need more validation with more studies. The climate factors of the top-five affected countries and three south Asian countries have considered in this study to have a real-time forecast and robust validation about the impact of climate variables.

**Methods:** The ARIMA model have included to model the univariate cumulative confirmed and death cases separately. The MLP, ELM and likelihood-based GLM count time series also considered as they consider the external variables as exogenous regressors. As the death count includes zero itself, zero-inflated count time series model has included instead of likelihood-based GLM. The better fitting of the ARIMA model will validate the under-whelm of meteorological factors was the initial hypothesis. The best model has identified through the application and comparison with the real data points.

**Results:** The results depict that there is an influence of meteorological variables like temperature and humidity mostly for all the selected countries cumulative confirm cases excluding Italy and Sri-Lanka. However, the best models for deaths count of each country also identify the impact of meteorological variables for each country.

**Conclusion:** The authors make the sixty days ahead forecast for each country which will be beneficial for the policymakers.

## 1. Introduction

The ongoing pandemic of novel coronavirus (COVID-19) became an acrimonious phobia for every citizen of the world as it already affects 212 countries and one international conveyance. The outbreak was primarily emerged in Wuhan (China) with severe and extensive contamination. However, a few weeks later, it rapidly spread all over the globe as the human-to-human transmission is an often event in this era of the global village. So far the data on the date of May 8, 2020 reveal that 4,000,282 persons have infected across the world with 275,323 death and 48,455 critical severe cases where the proportion of total cases and penalties for each million population is 513.2 and 35.3 percent (Worldometer, 2020; WHO, 2020). To control the outbreak of the pandemic, the identification and isolation of infected individuals or making social distance, are the most implemented methods since now. But the identification of the contact person is the most crucial part of this method; this is why the feasibility of making social distance home quarantine is also in a state of trepidation (Hellewell et al., 2020).

However, most of the countries try to control the outbreak by lockdowns of their cities and regions, countries in Europe still in a situation of nastiest as Europe became the epicenter of this pandemic. Until now, Italy, the USA, Spain, Germany, Iran, France, Switzerland, Iran, UK, and South Korea are the top affected countries after China, where most of them belong to the region of Europe. Among the top affected countries USA, Italy, Spain, France, and Iran are grappling the worst turmoil after the original one with an exponential increase of deaths. The novel coronavirus is similar to another epidemic named severe acute respiratory syndrome (SARS), but the total deaths and cases in COVID-19 already exceed multiple compared to the outbreaks of SARS in 2002-2003 (Lai et al., 2020; WHO, 2020). The outbreak of this pandemic influenced by several underlying factors. The debate of daily influences of the weather variables for the transmission of epidemic comes to an end after some recent studies. Moreover, the recent study indicates that wind speed, temperature, and relative humidity have a high correlation with the outbreak of the pandemic where another research specifies diurnal temperature is positively, and relative humidity is negatively associated with mortality or death count. Another study relates the climate variable with the doubling time where temperature show positive and evaporation show the inverse relationship (Oliveiros et al., 2020; Ma et al., 2020; Chen et al., 2020).

When it comes to the point of forecasting the count, several methods were already applied to have real-time forecasting. The short-term forecasting methods include generalized logistic growth model, Richard model, and sub-epidemic wave model applied to the data of 34 areas, including provinces, autonomous regions, and municipalities’ cumulative cases in a current study (Roosa et al., 2020). Another study uses the symmetric and Gauss function to identify and forecast the infected, suspected, and deaths in Hubei and China (Li et al., 2020). Moreover, the use of modified auto-encoders forecasting (Hu et al., 2020), several non-parametric model implementations in case of forecasting the spreading the pandemic at China, Italy, and France (Fanelli and Piazza, 2020), and the application of ARIMA base models also noticed to have the forecast of the epidemic (Benvenuto et al., 2020). All the mentioned study involves only the cumulative or infected case, new cases, and deaths of different province and country for forecasting, where most of the research goes through the univariate modeling approach. Examination influences are absent for meteorological variables, but there exist positive, as well as the negative association of it towards the outbreak, which already reported earlier. Moreover, the considered variables are integer or count in nature, but no approach of modeling with the basic count time series models noticed in the literature for COVID-19. Moreover, several usual machine learning and exogenous regressor-based prediction also not seen in the research for the COVID-19 pandemic analysis. This article aims to forecast the confirm, and the death count of the top outbreak or affected countries as well as some selected South Asian county with the consideration of meteorological factors of the individual state with the comparative study of conventional, machine learning, and count time series models.

The organization of this article as follows. Section 2 provides a detailed description of the data and research methods including an extreme machine learning algorithm and zero inflated count time series model. Section 3 demonstrates the results with relevant discussion about the significant findings. Finally, the summary of the key finding with concluding remarks are presented in Section 4.

## 2. Methodology

### 2.1. Data and Data Sources

The daily reported cumulative confirmed cases, and the number of deaths of the top five affected (China, USA, France, Germany, Italy, and Spain) as well as three South Asian countries (Pakistan, India, and Sri Lanka) along with the climate variables of each, are considered for this study. The climate factors consisting maximum, minimum, and average temperature, wind-speed (internal, guest, and average), wind-direction, perception (mm), total-cloud, wind-pressure, the humidity have selected with an average of available top affected province of the individual country along with cumulative confirm, recovered and deaths count. The data collected through two different R packages named Climate, and nCov2019, where both packages use different reliable and valid websites and institutional data. Readers suggest seeing the references to have a detailed idea about the sources of these data (Yu, 2020; Guidotti, 2020; Czernecki et al., 2020; Ogimet, 2020; danepubliczne.imgw.pl, 2020; Wyoming Weather Web, 2020). However, the missing data replaced by the average value of five previous and post data points from the missing data points. Factor type variables have replaced by the mode respectably. To make the comparison and model validation with the observed and forecasted data of different models, the data before the 27th of April have considered.

### 2.2. Methods

Data considered for this study indicates the univariate modeling approach of time series as we aim to forecast the cumulative infected cases, and deaths of selected countries. The autoregressive integrated moving average (ARIMA) is the most convenient model for predicting the univariate data. But in the case of predicting the count data, it’s often misleads. To have stationary data, usually, the difference or transformation of data is required for the ARIMA model, which may invalidate the uniqueness of count data as it holds only the integer value. Apart from these tricky things, ARIMA still considered for count time series forecasting as related literature already mentioned in the prior. However, the prediction based on the machine learning algorithm also enriched the research in recent times, but several studies report the inadequacy of performing for a small volume of data. Moreover, machine learning models do not have any specialization for count data as they only require the quantitative structure of the data.

However, the concern about the influence of the climate variable is the focal point for this study. Several ML time series models give the opportunity of univariate forecasting with the inclusion of exogenous regressors. Among the machine learning algorithm, Multi-layer-perceptron (MLP) and Extreme learning machine (ELM) algorithm for time series forecasting have considered for making the comparison to other models. It needs to mention that the deep learning forecasting algorithms such as Long-Short-Term-Memory (LSTM) neural network, also applied to forecast the epidemic. Still, the considered data for the individual country are small in volume, and the results also show a large quantity of error for the pilot study, which has no way to engage with included models. In contrast, the conventional Poisson and Negative-Binomial distribution base count time series models may be the most accurate way of presenting the analysis and forecasting as the projected data consist of the integer value. Besides, among the bunch of count time series models, likelihood-based generalized linear models with Poisson and Negative-Binomial conditional distribution is considered for this study, as it found usable in the literature compared to some recurrently used count time series model (Liboschik et al., 2017). Conversely, handling the zero in count data is also an important task. The cumulative death data found for this study consist of many zero as initial days of pandemic do not report any death at most of the country. Thus, data-driven methods of count time series or zero-inflated model also included in the study for giving access to the zero in the forecasting.

### 2.3 Mathematical Illustration

The ARIMA model is probably the most popular technique for univariate forecasting; hence the mathematical illustration of ARIMA is not given in the following section. To get more details about the modeling and precise forms of ARIMA, a reader suggests seeing the referred book (Shumway and Stoffer, 2000). However, the mathematical demonstration of the considered model or basic idea of included models have discussed in the following section; inferential and precise form is available on referred books and journals.

#### 2.3.1 Multi-Layer-Perceptron (MLP) with Exogenous Regressor

To have accurate time series prediction with specifying artificial neural network architecture is being too hassle-free after the proposed methodology of Crone and Kourentzes (Crone and Kourentzes, 2010; Kourentzes et al., 2014). An entirely data-driven technique of automatic network specification from the pattern of data and time-frequency have specified with the combination of filter, the transformation of the explanatory variable, feature evaluation, and specification of multilayer perceptron. The architecture with independent and dependent variable determine the relationship 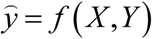 with predicted value 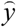. For time series forecasting, a feedforward NN is built with 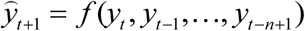 input vector with n lagged *y_t_*_−_*_n_* dependent variable where the NN is constructed a functional form 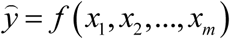 for mexplanatory variable and *x_m_* metric. The authors try to develop a model by using the analogy of auto-arima where the limit of order is limited to one to fourteen. The single output MLP function can be written as,

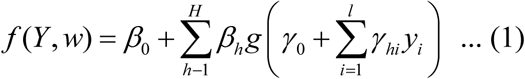

where, vector lagged observation with *n = p* lag and *n = I* input unit from n preceding point *t*,*t*−1,*t*−2*,…,t*−*n +*1, with Bias *β*_0_ *and γ*_0_*_i_* of each node the weights for hidden and output layer is *w=(β,γ*), *β=[β*_1_*,β*_2_*,…,β_H_*] and *γ = [γ*_11_*,γ*_12_*,…,γ*_21_*,…,γ_hI_*] respectively where number of input and hidden units in the network specified by *I and H specify*. To select the feature authors, suggest a combined filter with Wrapper approach for time series prediction. To have an automatic feature evaluation, Box-Jenkins methodology with the calculation of minimum Euclidean distance or with the identification and fitting of seasonality for minimum distance have used. The transformation and automatic feature construction with the identification of accuracy and robustness have done with the INF algorithm. To have the brief idea reader suggests seeing the referred articles and book of Nikolaos Kourentzes and others (Crone and Kourentzes, 2010; Kourentzes et al., 2014; Ord et al., 2017).

#### 2.3.3 Extreme Machine Learning Algorithm

The conventional feedforward neural network learning speed is slower because of the slow gradient-based algorithm and the iterative algorithm of tuning the parameter. Huang and others proposed an extreme learning machine (ELM) by randomly chosen hidden nodes and output weight with single hidden layer feedforward neural networks (SLFNs) to skip these problems (Huang et al., 2006). They suggest using the minimum norm least-squares solution of SLFNs instead of the conventional gradient-based solution. However, the proposed algorithm is of an extreme machine learning technique which is briefly discussed as bellow,

With hidden node number *Ñ* and activation function *g(x)* given training set,

**Step 1:** Input Weight *w_i_* and bias *b_i_* is assigned randomly with *i* = 1,…,*Ñ*.
**Step 2:** Calculation of output matrix of hidden layer H.
**Step 3:** Calculation of output weight *β =* H^*^T with T = [*t*_1_,…,*t_N_*]*^T^*.

Moreover, the ensemble operators used in both algorithms consisting mean, median, and mode ensemble have used as followed by Nikolaos Kourentzes and others (Kourentzes et al., 2014). To have the detailed discussion about the mentioned algorithm reader should go through the referred articles (Huang et al., 2006). The estimation and forecasting of ELM and MLP algorithms have determined through a newly introduced R package named *nnfor* (Kourentzes, 2019).

#### 2.3.4 Likelihood-Based Generalized Linear count time series Model

The GLM with likelihood-based is analogous to the generalized autoregressive conditional heteroscedasticity (GARCH). This procedure involves the Poisson and Negative-Binomial distribution as conditional distribution with logarithmic and identity link function where the INGARCH model can consider as a particular case. The model general functional form can write as,

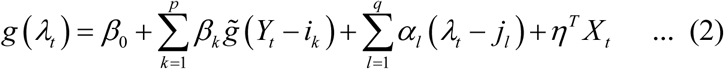

where the conditional mean 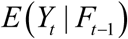 of the count time series such that *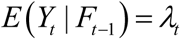* With joint process 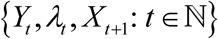 up to *F_t_* history with link *g*: ℝ^+^→ ℝ;, transformation function *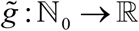*, parameter vector *η* =(*η*_1_,…,*η_r_*) and linear predictor *v_t_ = g*(*λ_t_*). The set 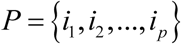 and 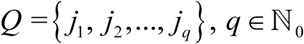, *q* ∈ ℕℕ_0_ allow the regression on arbitrary past observation which enables to regress on lagged observation *λ_t−j_*_1_*, λ_t−j_*_2,_*…,λ_t−jq_*. Model (2) can be considered for Fokianos and Tjøstheim delivered log-linear model with link function 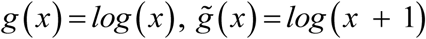 with 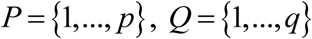 and linear predictor *v_t_ = log* (*λ_t_*). The model can be written as,

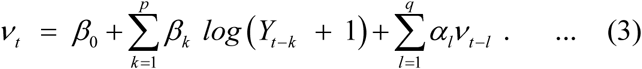

By holding 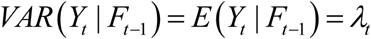 and 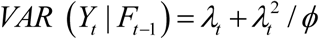, for Poisson and Negative-Binomial distribution assumption 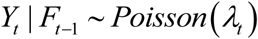 and *Y_t_* | *F_t_*_−1_ ~ NegBin(*λ_t_*,*ϕ*) respectively the conditional distribution can be written as following,

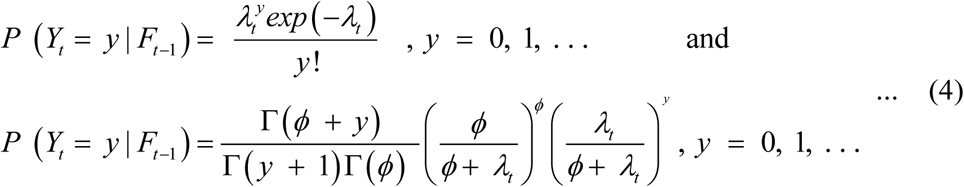

where dispersion parameter *ϕ* ∈ (0, ∞) and conditional variance 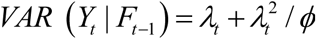.

This model includes internal covariates effect by the dynamic propagation to future observation by regression of past observation and past conditional means. The external covariates effect included intervention effects followed by the theory of Liboshchick and others (Liboschik et al., 2016; Karimuzzaman et al., 2020; Rahman and Harding, 2017). However, both internal and external covariates effect is allowed by the following generalization of the model (eq 2) as,

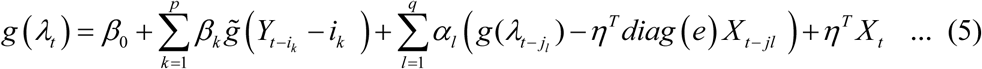

The estimation and inferences of the described model have made with the theory of quasi conditional maximum likelihood estimation with quasi Poisson assumption; otherwise, it obtains an ordinary maximum likelihood estimator. However, the detail estimation and inference procedure with the prediction algorithm, the inclusion of intervention analysis, and model assessment have explained in the refereed journal of Liboschik and others (Liboschik et al., 2017; Liboschik et al., 2016). The computation and application of the mentioned model are available on R package tscount (Liboschik et al., 2017), and readers can also suggest to see the details applications and distinction with other open packages (Liboschik et al., 2017).

#### 2.3.5 Zero Inflated Count Time Series Model (ZIM)

The zero-inflated version of Poisson and Negative-Binomial distribution is replaced instead of ordinary distribution in the count time series model to give an excess of zero. This version of models allows the mixture of singular distribution in zero and the conventional Poisson and negative binomial distribution with *W_t_* and 1 − *W_t_* probability respectively. The idea of these types of models are available in the literature since the application count regression model, but the proposed data driven model of Yang, Zamba, and Canavag (Yang et al., 2013) gives new influence over the count time series modeling. Proposed method allows the probability *(W_t_*) with a time varying GLM logit link and the conditional mean (*λ_t_*) is model through logistic regression model which also vary over time. However, the parameter of the ZIM fitted model with the estimation of the EM algorithm. This model also includes an extension of state-space models (Yang et al., 2015). However, the proposed model may name as Poisson autoregressive model in the partial likelihood framework where a Markov regression model may develop for count time series with an excess of zeros. Among the several mathematical demonstrations, we include only the fundamental theoretical part, for details reader may review the referred article.

Let 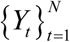 count time series follow ZIP (λ_t_, ω_t_) with Probability mass function,

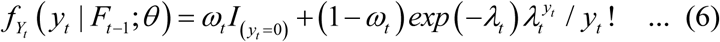

where *F_t−_*_1_ work as filtration parameter. The cumulative distribution function can be written as

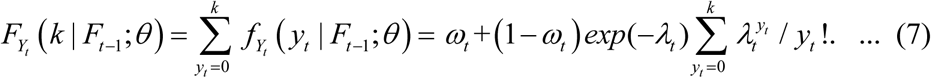

With non-negative integer *K*, cumulative function *Y_t_* | *F_t_*_−1_, mean *E*(*Y_t_* | *F_t_*_−1_;*θ*) = *λ_t_* (1*−ω_t_*) and variance *Var(Y_t_* | *F_t_*_−1_;θ) = *λ_t_* (1*−ω_t_*)(1 *+ λ_t_ω_t_*). ZIP distribution can work for both over-dispersion and zero inflation, since the variance is always greater than mean. However, the ZIP autoregression can be written as,

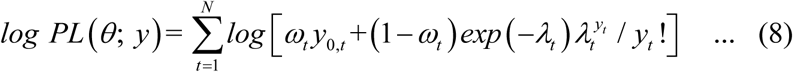

With partial likelihood 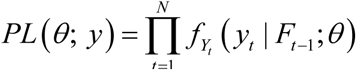, and parameter *λ_t_* and *ω_t_*. The parameter can be defined as 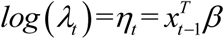 and 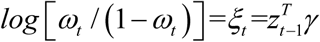 where 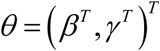 and 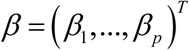 and 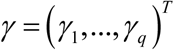. However, the application and computing were done through the R Package ZIM (Yang et al., 2014).

## 3. Results and Discussion

The pandemic is now in a horrific position as it spread over the 212 countries over the world. Among them, the top fifteen countries break the record every day as of their previous day death number (Fig. 1). The concentration of this study was to identify the best forecasting model among the selected models with the inclusion of climate variables along with recovered as exogenous regressor for death and infected count. In other words, the robust validation of ongoing debate about the effect of climate on virus spreading has made through the inclusion of climate factors. If there is an existence of better forecast from those models, which includes the meteorological variables as an exogenous regressor (MLP, ELM, Likelihood-Based GLM, and Zero-inflated models), that may indicate the validation of the effects of meteorological variables. The univariate ARIMA model has considered for the comparison with no inclusion of external regressors. So, if there is any evidence of better forecasts through the ARIMA model, that will nullify the effects of meteorological variables. However, the top five-country of this global pandemic and three selected South Asian countries have studied for the comparison and contrast among the applied algorithms.

**Figure 1:**
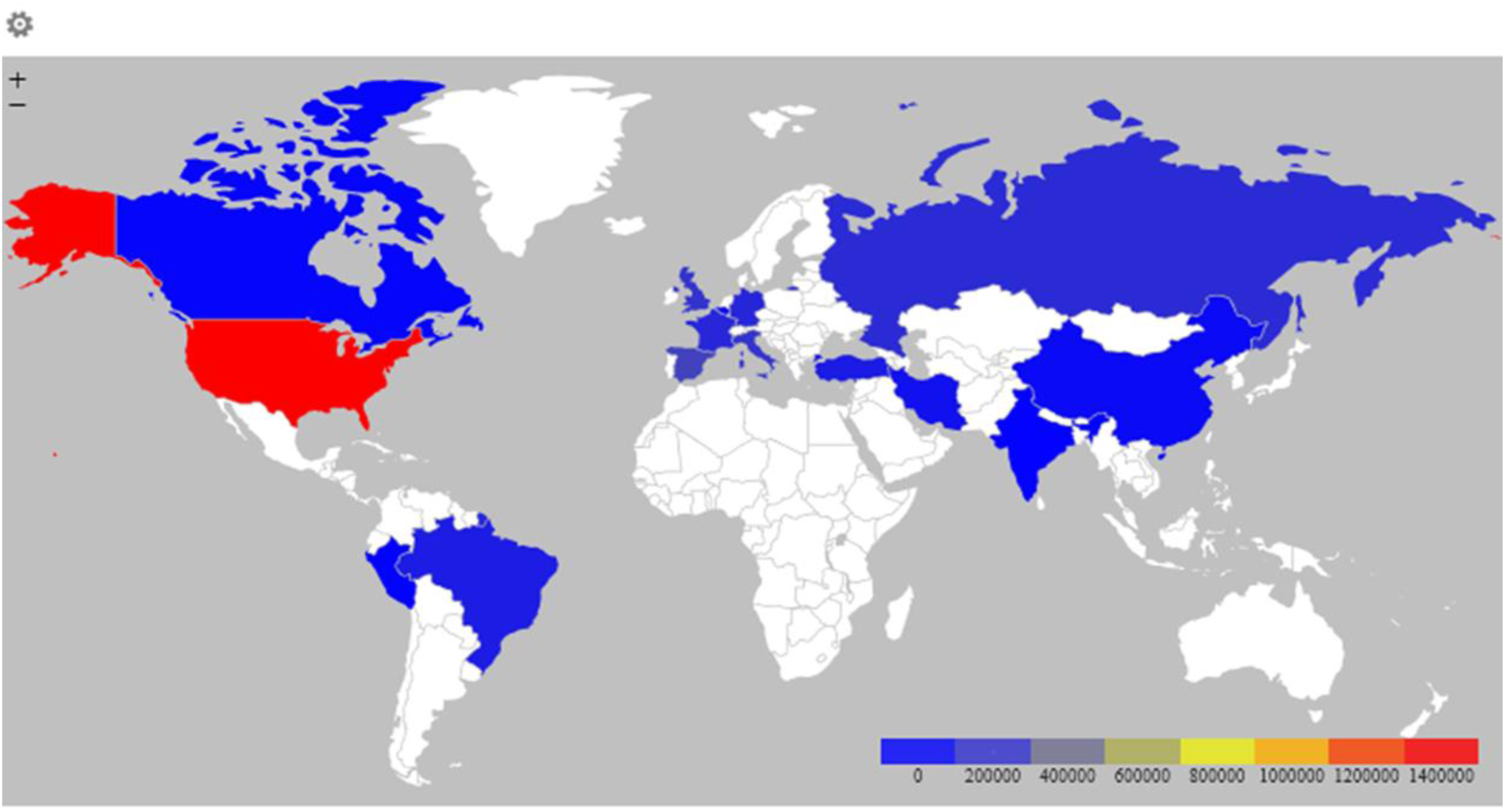
Top Fifteen COVID-19 Affected Country

These aforementioned models were applied for each of the countries individually; hence the comparison was made with the conventional models, machine learning algorithm and likelihood base generalized linear model distinctly. The model accuracy and model selection criteria’s have calculated and showed for both the cumulative cases and deaths (see, Table 1 and Table 2).

**Table 1:** MSE of Machine Algorithms for Cumulative cases and Deaths

| Country | Model | Cumulative Confirm | Cumulative Death |
| --- | --- | --- | --- |
| China | MLP | 6369.5048 | 1.039 |
|  | ELM | 317043.6732 | 276.1357 |
| USA | MLP | 4433.5751 | 1.0869 |
|  | ELM | 30016.2018 | 1.4197 |
| France | MLP | 168.097 | 38027.4188 |
|  | ELM | 0.108 | 233.3686 |
| Germany | MLP | 27027.2672 | 0.9565 |
|  | ELM | 83998.869 | 3.0211 |
| Italy | MLP | 2738.8021 | 638.392 |
|  | ELM | 91725.1556 | 1206.7364 |
| Spain | MLP | 19873.0188 | 34.3813 |
|  | ELM | 28899.0753 | 659.3208 |
| Pakistan | MLP | 54.2123 | 0.0327 |
|  | ELM | 740.7721 | 2.2348 |
| India | MLP | 177.905 | 0.0598 |
|  | ELM | 258.3877 | 2.28103 |
| Sri-Lanka | MLP | 0.5117 | 0.0161 |
|  | ELM | 17.413 | 2.28103 |

**Table 2:** Model-Selection Criteria of Cumulative Confirm Cases for Likelihood Based-GLM

| Country | Model | AIC | BIC | QIC |
| --- | --- | --- | --- | --- |
| China | Poisson | 3257.57 | 3301.858 | 3257.57 |
|  | Negative-Binomial | 1403.943 | 1450.836 | 145632.1 |
| USA | Poisson | 945.2496 | 980.8534 | 945.2496 |
|  | Negative-Binomial | 186.988 | 222.5926 | 186.988 |
| France | Poisson | 1336.849 | 1345.607 | 430090 |
|  | Negative-Binomial | 1361.132 | 1369.891 | 571581.3 |
| Germany | Poisson | 870.4162 | 904.9583 | 869.7685 |
|  | Negative-Binomial | 736.0338 | 772.7348 | 38397.21 |
| Italy | Poisson | 930.1754 | 964.1321 | 930.1052 |
|  | Negative-Binomial | 860.6978 | 896.8298 | 58441.53 |
| Spain | Poisson | 1437.995 | 1464.436 | 1435.877 |
|  | Negative-Binomial | 416.5113 | 444.5076 | 17479.53 |
| Pakistan | Poisson | 68727.47 | 68763.6 | 1435.877 |
|  | Negative-Binomial | 416.5113 | 444.5076 | 17479.53 |
| India | Poisson | 1188.574 | 1224.736 | 1184.104 |
|  | Negative-Binomial | 736.0338 | 772.730338 | 38397.21 |
| Sri Lanka | Poisson | 262.862 | 299.5631 | 281.6033 |
|  | Negative-Binomial | 197.568 | 135.792 | 145.077 |

To have an initial idea about the model fitting of the MLP and ELM models, Mean Sum Squares of Error (MSE) has calculated. Conversely, among the Poisson and Negative-Binomial likelihood-based GLM, the fundamental distinction drawn through the reported Akaike Information Criteria (AIC) and Bayesian Information Criteria (BIC) along with Quasi-likelihood Information Criterion (QIC). However, the model accuracy of the ARIMA model also reported where model accuracy for both cumulative death and confirmed cases have indicated for each country (see, Table 3). The MLP, ELM, ARIMA, and Likelihood base GLM count time series have applied for cumulative confirm claims. Since the cumulative death counts consist of zero itself, the zero-inflated model has used instead of Likelihood base GLM to forecast cumulative death count. Among the machine learning algorithms, MLP seems to have better forecasting accuracy as MLP shows lower MSE for both confirm and death cases except France (see, Table 1).

**Table 3:** ARIMA Order and Accuracy

| Country | ARIMA (Order) | RMSE | MPE | MAPE |
| --- | --- | --- | --- | --- |
| China | Confirm (1,2,0) | 1430.408 | 2.142756 | 6.201592 |
|  | Death (0,2,1) | 15.62574 | 3.502292 | 8.57898 |
| USA | Confirm (2,2,0) | 292.3055 | 1.127264 | 15.29551 |
|  | Death (2,2,2) | 1.87418 | 5.935086 | 12.8242 |
| France | Confirm (0,2,4) | 101.989 | 1.982609 | 11.21796 |
|  | Death (0,2,3) | 9.841609 | 3.670882 | 21.23384 |
| Germany | *Confirm (0,2,2)* | 340.2773 | 2.059632 | 10.56923 |
|  | Death (0,2,4) | 1.837792 | 12.3112 | 27.35068 |
| Italy | *Confirm (3,2,2)* | 639.9403 | 4.096929 | 7.783249 |
|  | Death (0,2,2) | 61.34247 | 5.737476 | 14.62763 |
| Spain | *Confirm (0,2,0)* | 705.1241 | 4.47149 | 10.14358 |
|  | Death (2,2,2) | 36.40452 | 8.634018 | 12.08562 |
| Pakistan | Confirm (0,2,1) | 36.73136 | 3.620884 | 13.25082 |
|  | Death (0,2,2) | 1.670163 | 3.555396 | 29.06964 |
| India | Confirm (2,2,0) | 7.323552 | 2.27114 | 8.645043 |
|  | Death (0,2,2) | 0.238028 | 13.61276 | 23.78294 |
| Sri Lanka | *Confirm (2,2,3)* | 7.323552 | 2.27114 | 8.645043 |
|  | Death (0,2,2) | 0.1258904 | 32.13213 | 37.48764 |

Machine learning (ML) is a procedure of learning from the data, and the learning scheme makes the difference between the algorithms. The distinction of algorithms through MSE or any other accuracy measurement may fail to identify the real one, as ML is a learning procedure from the data. Hence, there is a prerequisite for further demonstration for detecting the best algorithm. Graphical representation and comparison of predicted data towards the observed value may give better shades on discovering the best algorithm. Moreover, the inclusion of different models has a diverse way of handling the count time-series facts. In the likelihood-based generalized linear count time series model, Poisson and Negative-Binomial distribution with link function considered as conditional distribution. Hereafter, the best-fitted model between the Poisson and Negative Binomial has chosen through the AIC, BIC, and QIC, where every state shows low values for the negative binomial distribution base generalized linear model (see, Table 2).

The graphical comparison among Observed, ARIMA, MLP, ELM, and Likelihood-based Generalized count time series model have drawn to have an ultimate better-fitted model for the cumulative confirm cases. Initially, ten days forecast have drawn for making the comparison with a real one (named as observed) for all the countries according to the available number of data points. However, according to the graphical deduction, likelihood-based generalized count time series have shown to have forecast better except Italy, Spain, and Sri-Lanka. The ARIMA, and ELM considered to be as better forecasting algorithm for Italy and Spain. Sri-Lankan cumulative confirm cases pattern cannot explained through any of the applied models. Thus, MLP have considered for the further forecasting as it shown a more reliable than others (see in the Fig. 2).

**Figure 2:**
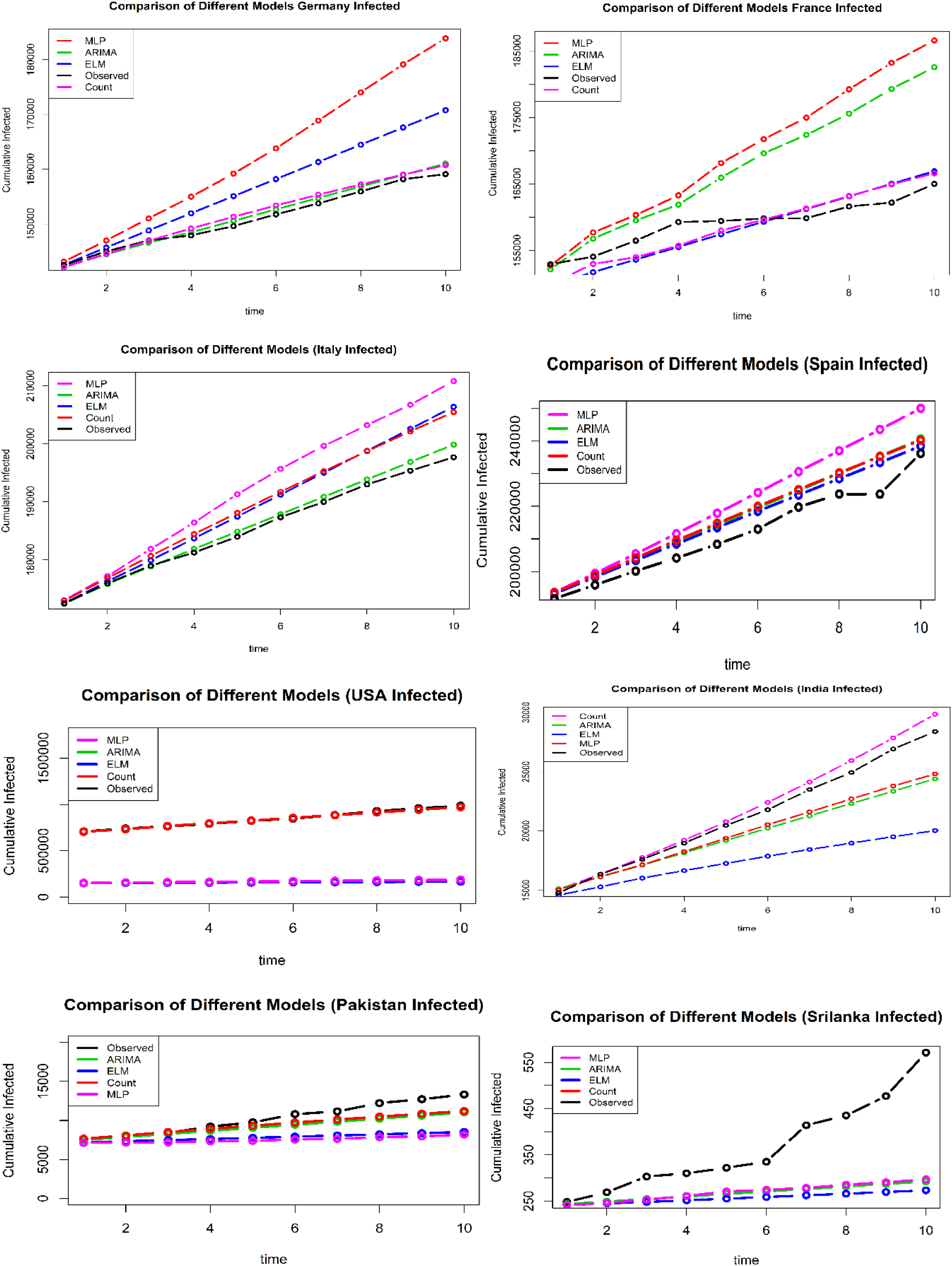
Forecast Comparison of Cumulative Confirm

Similarly, the graphical comparison also illustrates for the death forecasting where most of the death cases forecasting show the reliability from the machine learning algorithm. The ELM is as appropriate algorithm for France, Germany, and Spain, where MLP seems to give better forecast for India, and Pakistan. The popular ARIMA model seems to fit well for Sri-Lanka, Italy, and USA death forecast (see in, Fig. 3).

**Figure 3:**
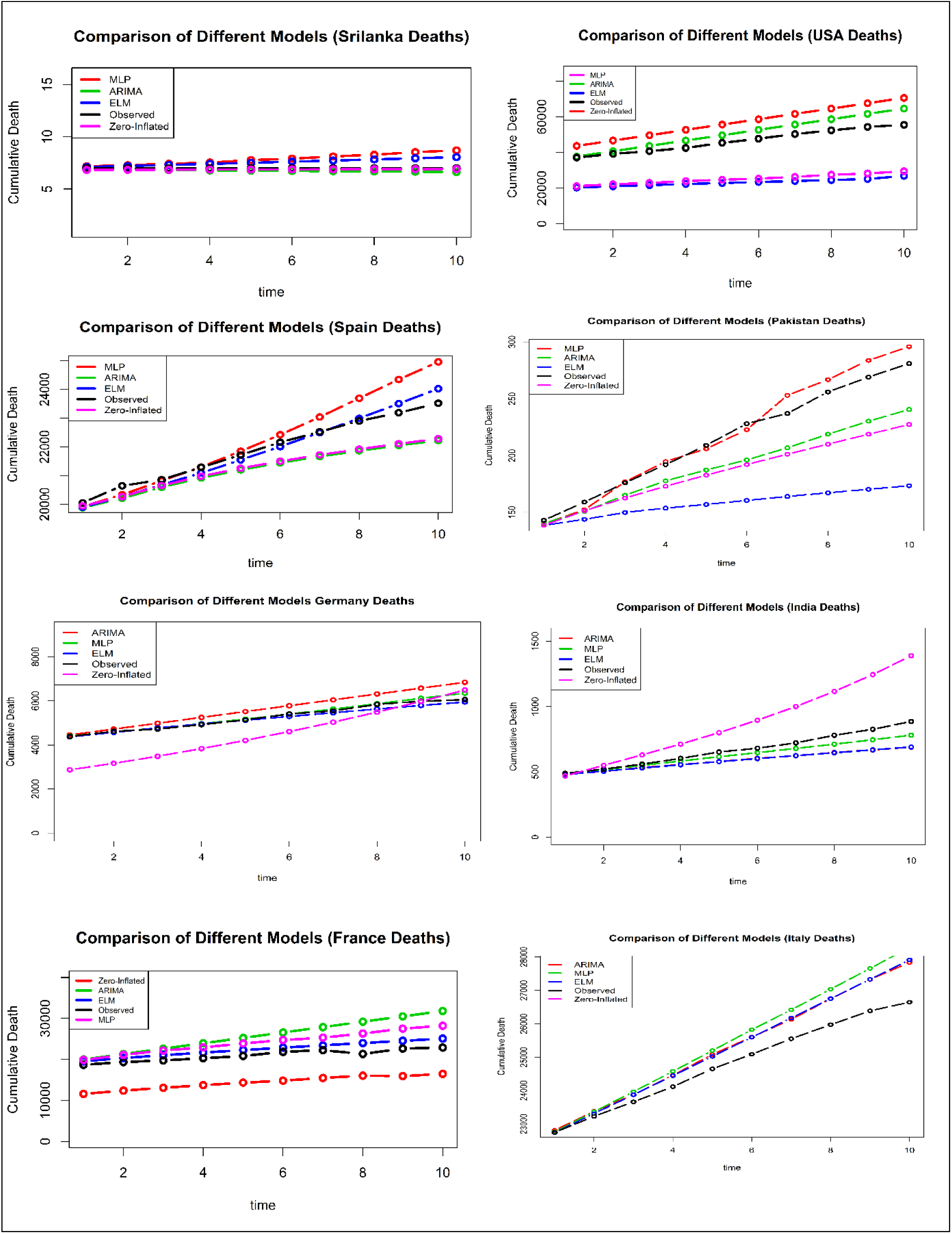
Forecast Comparison of Cumulative Death

The ARIMA model with the log transformation makes the data stationary as an augmented Dickey-Fuller (ADF) test for each of the country gives significant p-values at 5% level of significance. However, the forecasting accuracy, along with their autoregressive and moving average order of each state also displayed in this article (Table 3).

Nevertheless, neural network structure with hidden layer and nodes of individual fitted machine learning algorithm presented throw the respective figures (see, Fig. 4).

**Figure 4:**
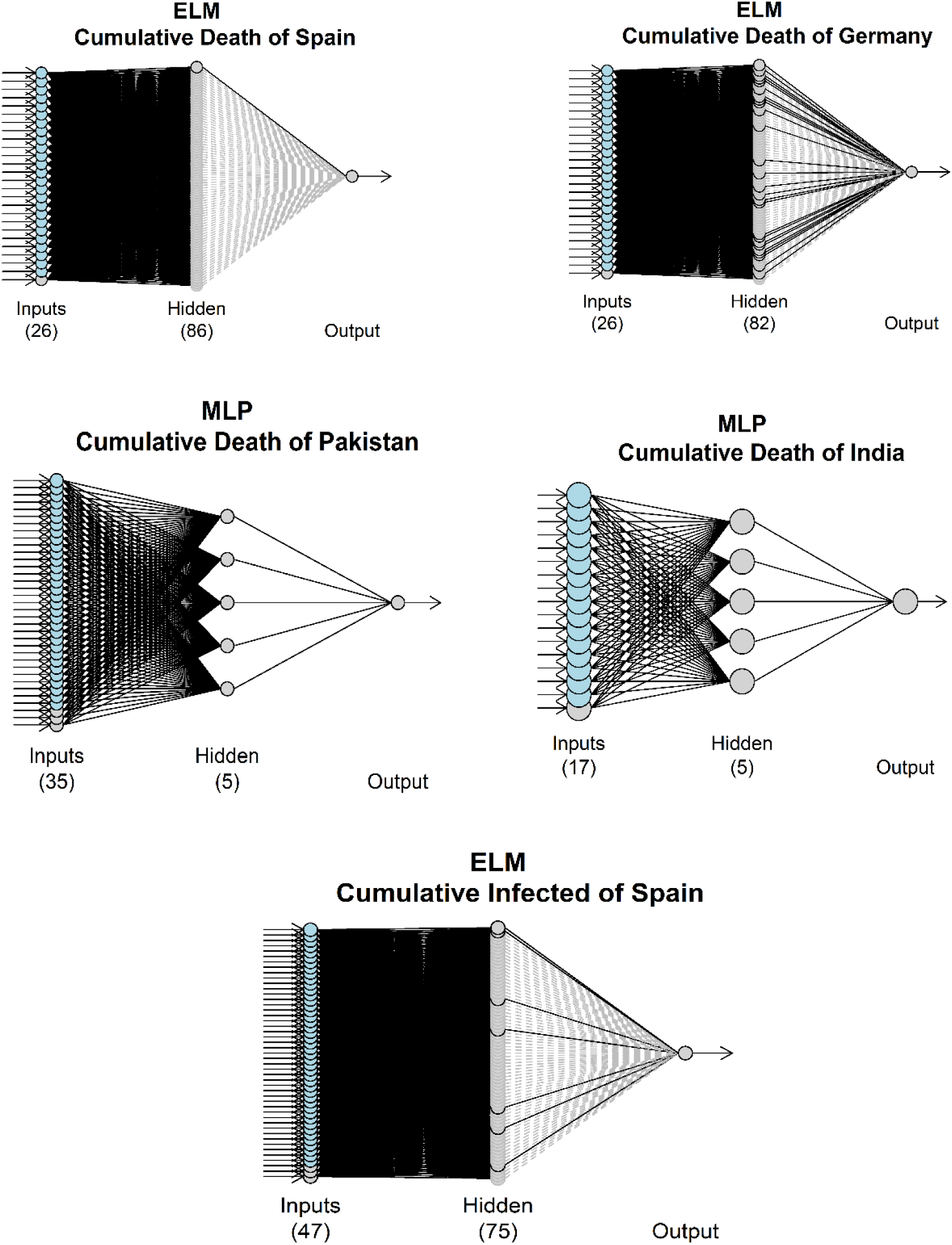
Neural Network Structure for Cumulative Death and Infected Cases

It needs to mention that, zero-inflated count time series regression model has fitted for every country with the overdispersion test and EM algorithm base parameter estimation. But it failed to comes in its best way as compared to others. However, the model parameters, over-dispersion test results, as well as AIC and BIC, showed for each country (Table 4).

**Table 4:** Zero Inflated Count Time Series Model Parameter

| *Country* | *Overdispersion*<br>*score- test (P-value)* | *Number of EM-NB*<br>*Iteration* | *AIC* | *BIC* |
| --- | --- | --- | --- | --- |
| China | 2.22e-16 | 11 | 4797.792 | 4839.475 |
| France | 0.92885 | 11 | 303.2677 | 337.6987 |
| India | 0.96855 | 25 | 137.3223 | 171.3564 |
| Italy | 2.22e-16 | 11 | 3069.84 | 3100.109 |
| Pakistan | 7.3344e-16 | 11 | 158.6184 | 161.7291 |
| Spain | 2.22e-16 | 11 | 1667.176 | 1701.3 |
| USA | 0.99671 | 12 | 230.4939 | 264.0034 |
| China | 2.25e-16 | 15 | 1237.23 | 1549.673 |
| Sri Lanka | 0.88525 | 3 | 12.94 | 18.50 |

There is an extensive influence of climate variables as the analysis and illustration shows that the regressor base count time series model gives the better forecast model for every selected country excluding Italy. The death forecasting stimulation also illustrate the analogous consequence as exogenous regressor base machine learning algorithm appears to demonstrate the better forecast. Italy seems to fit better with ARIMA model for both cases, which depict the inexistence of the influences of climate variables. Sri-Lanka shows a better forecast for death with ARIMA model, where none of the applied models illustrate the real scenario of infected cases for Sri-Lanka that indicate the inexistence of the influences of climate variables for Sri-Lanka. Finally, the sixty days forecast of each of the cumulative confirm cases and deaths have estimated for selected countries using respective selected models and algorithms.

Furthermore, maximum likelihood base count time series have selected for all the countries excluding Italy and Sri-Lanka to forecast the cumulative confirm cases. Conversely, ELM for Germany, France, and Spain; MLP for India and Pakistan; and ARIMA for others have used for forecasting the deaths. Results demonstrate that after 27th April 2020 the next thirty days France, Germany, India, Italy, Pakistan, Spain, Sri-Lanka, and USA will have respectively 199317, 174159, 136471, 259844, 350896, 19839, and 1306159 individuals as infected. And after the next thirty days, the number of infected people will be 230867, 177194, 638604, 324499, 20601, 462152, 39679, and 1400845 (see in, Fig. 5).

**Figure 5:**
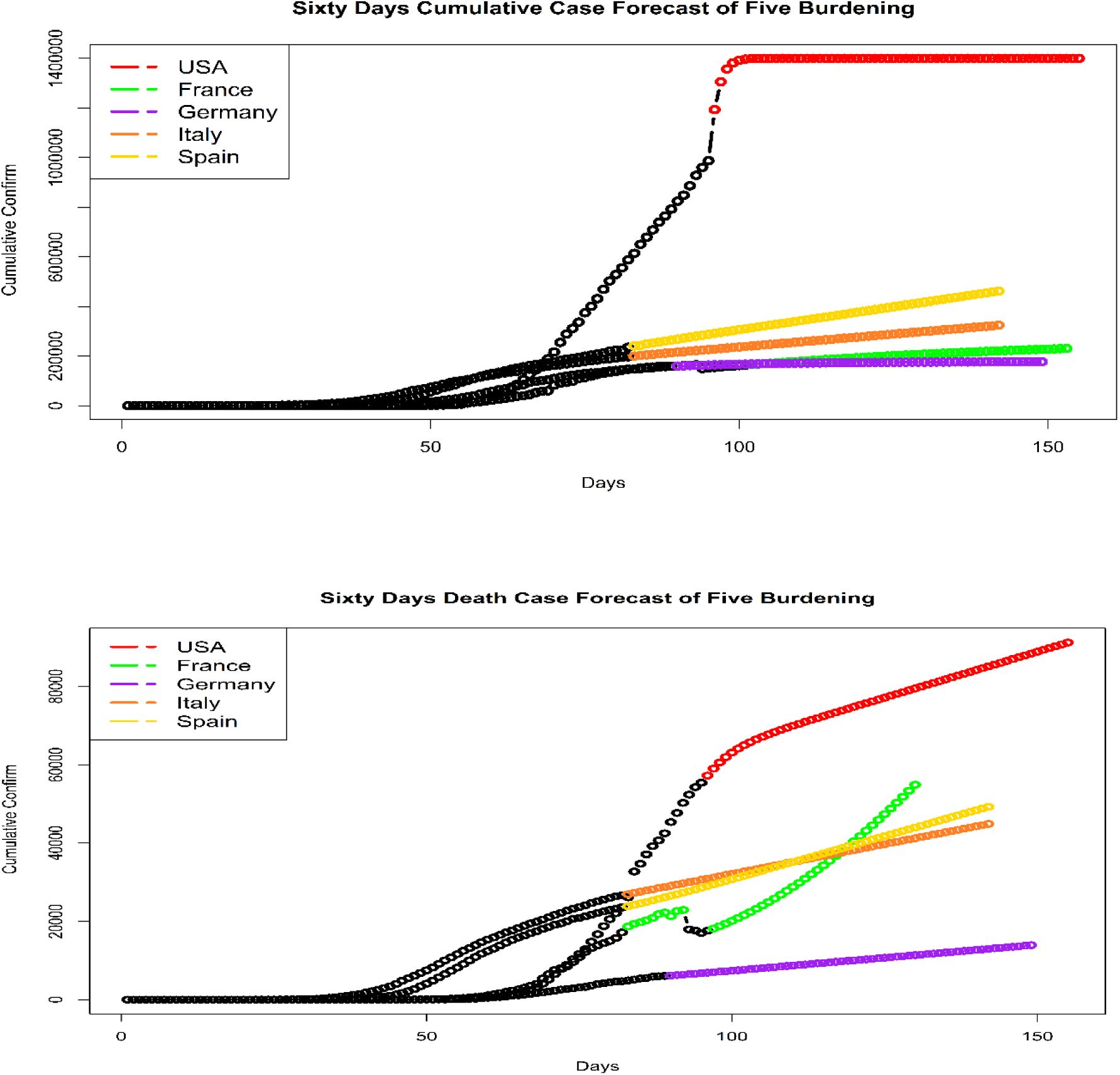
Top Five Affected Country Cumulative Confirm and Death Forecast

The authors also forecast the number of deaths of France, Germany, India, Italy, Pakistan, USA, Sri-Lanka, and Spain. The results depict that after 27th April 2020 the next thirty days death forecast will be 28797, 9906, 2699, 35768, 734, 77134, 22, and 36046. Finally, the sixty days deaths forecast is 54899, 13896, 4520, 44877, 1188, 91203, 49, and 49246 for France, Germany, India, Italy, Pakistan, USA, Sri-Lanka, and Spain respectively (see in Fig. 6).

**Figure 6:**
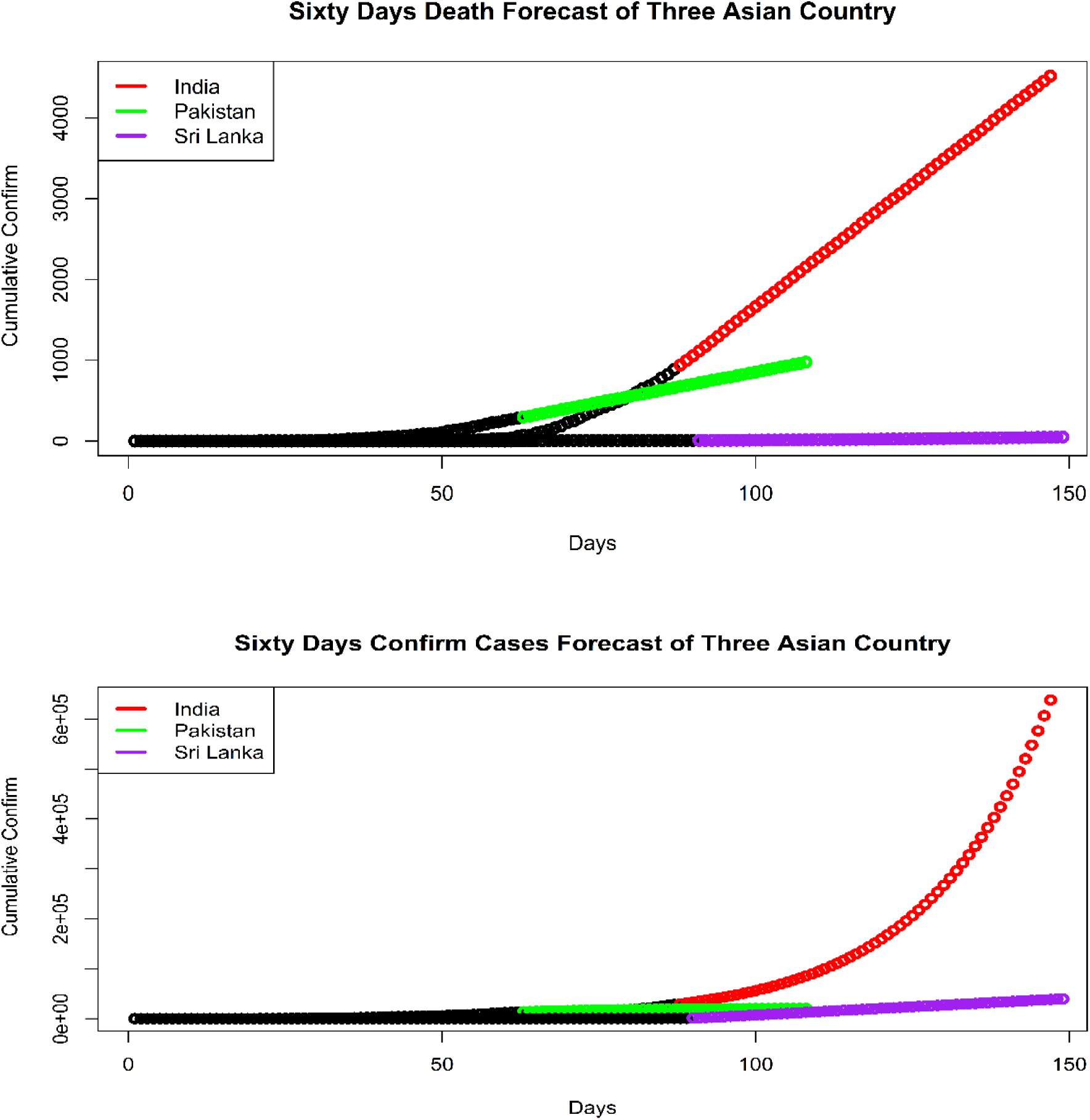
Three Selected Asian Country Cumulative Confirm and Death forecast.

## 4. Summary and Conclusion

Forecasting based on several epidemiological theories and methods has seen in the existing literature for COVID-19. Meanwhile, most of the studies used the well-known ARIMA model to forecast the COVID-19 cases. The cumulative confirm cases and number of deaths are integer-valued itself, which indicate the modelling should have done through the count time series approach with the inclusion of count distribution such as Poisson and Negative Binomial. Death count consist lots of zero itself, so the model with excess of zero is more appropriate here. Conversely, machine learning models handles the numerical data and do not take consideration the data type. From the starting of the pandemic, there is a debate about the influence of climate variables on spreading the COVID-19. Hence, the meteorological variables were included in this study as an exogenous regressor or covariates to have partial validation. The authors also include the univariate ARIMA model for comparing with other regressor base models. If the ARIMA model gives better forecast that will nullify the influences of the meteorological variable. This study considers the top five affected country and three south Asian countries for modelling purpose. Hence, the comparison was done through several calculative as well as graphical methods and found that there is an influence of meteorological factors for all the countries excluding Italy and Sri-Lanka to increase the infected cases. However, the best models for deaths count of each country also identify the meteorological impact for each country.

Furthermore, the forecasting of cumulative affected cases has done after the comparison among ARIMA, ELM, MLP, and Likelihood-based GLM, which predict a total of sixty days the possible number of cumulative confirmed cases. Results have demonstrated that after 27th April 2020 the next thirty days France, Germany, India, Italy, Pakistan, Spain, Sri-Lanka, and USA will have respectively 199317, 174159, 136471, 259844, 350896, 19839, and 1306159 individuals as infected. And after the next thirty days, the number of infected people will be 230867, 177194, 638604, 324499, 20601, 462152, 39679, and 1400845 (see in, Fig. 5). Similarly, the death forecasts of France, Germany, India, Italy, Pakistan, USA, Sri-Lanka, and Spain depict that after 27th April 2020 the next thirty days death forecast will be 28797, 9906, 2699, 35768, 734, 77134, 22, and 36046. Finally, the sixty days deaths forecast is 54899, 13896, 4520, 44877, 1188, 91203, 49, and 49246 for France, Germany, India, Italy, Pakistan, USA, Sri-Lanka, and Spain respectively. To finish, these forecasted results for each country would assist the policymakers in each country to make informed decision to control the risks. Given that the COVID-15 pandemic epicenters may change from western countries to some Asian and African countries, our future research will focus on more countries in those domains by using other factors such as geospatial and community specific factors.

## Data Availability

All data will be available upon request and/or in an URL.

## Contributions

Study design: MK, SA, MMH, AR; Data curation: MK, SA; Methodology: MMH, AR; Data analysis: MK, SA, MMH; Writing: AH, MK, MMH, AR. Overall supervision: AR. All authors read and approved the final version of the manuscript.

## Ethical approval

No ethical approval is required as this study based on aggregated COVID-19 surveillance data.

## Acknowledgements

The authors are greatful to the Editors for checking the manuscript carefully and providing comments which were useful to improve the manuscript.

